# Diffusion basis spectrum imaging in post-hemorrhagic hydrocephalus of prematurity

**DOI:** 10.1101/2021.01.12.21249706

**Authors:** Albert M. Isaacs, Jeffrey J. Neil, James P. McAllister, Sonika Dahiya, Leandro Castaneyra-Ruiz, Harri Merisaari, Haley E. Botteron, Dimitrios Alexopoulous, Ajit George, Sun Peng, Diego Morales, Yan Yan, Sheng-Kwei Song, David D. Limbrick, Christopher D. Smyser

## Abstract

**Objective:** The debilitating neurological deficits of neonatal post-hemorrhagic hydrocephalus (PHH) have been linked to periventricular white matter injury. To improve understanding of the deleterious mechanisms underlying PHH-related brain injury, this study applied diffusion basis spectrum imaging (DBSI) for the first time in neonates, modeling white matter fibers to assess axonal and myelin integrity, fiber density, and extra-fiber pathologies including cellularity, edema, and inflammation. The objectives of the study were to characterize DBSI measures in key periventricular white matter tracts of PHH infants, associate those diffusion measures with ventricular size, and utilize postmortem white matter histology to compare with the MRI findings.

**Method:** A prospective cohort of very preterm infants (n=95) underwent MRI at term equivalent age, of which 68 were controls (VPT group), 15 had high-grade intraventricular hemorrhage without hydrocephalus (IVH group), and 12 had PHH (PHH group). DBSI metrics extracted from manually segmented corpus callosum (CC), corticospinal tracts (CST), and optic radiations (OPRA) included fiber level axial diffusivity (FAD), fiber radial diffusivity (FRD), fiber fractional anisotropy (FFA), fiber fraction (FF), restricted fractions (RF), and non-restricted fractions (NRF). All measures were contrasted across groups and correlated with frontal occipital horn ratio (FOHR), a measure of ventricular size. Postmortem immunohistochemistry was performed on the CC of 10 preterm infants (five VPT, three IVH, and two PHH) and two full-term infants who died from non-neurologic causes assessing white matter intra- and extra-fiber pathologies, as well as the integrity of the adjoining ventricular and subventricular zones.

**Results:** Except for FF in the CC, there were no differences in all measures between IVH and VPT infants. In the unmyelinated CC, PHH had the lowest FF, FAD, and FFA and the highest RF. In the CC, FOHR related negatively with FAD, FFA, and FF and positively with RF. In the myelinated CST, PHH had the lowest FAD, FFA, and FF and the highest FRD and RF. FOHR related negatively to FAD and FFA and positively with NRF and FRD. In the OPRA, PHH was associated with the lowest FF and the highest RF, NRF, and FAD. FOHR related positively with FAD and NRF and negatively with FF. On postmortem tissues, PHH was associated with the highest white matter cellularity counts, variable amounts of cytoplasmic vacuolation, and the lowest synaptophysin marker intensity. The adjoining ventricular and subventricular zones in PHH had poor cytoarchitecture on H&E staining and relatively increased expression of GFAP and IBA1.

**Conclusions:** This initial utilization of DBSI to investigate neonatal brain development and injury demonstrated that PHH was associated with diffuse periventricular white matter injury, with tract-specific microstructural patterns and severity of axonal injury, myelin injury, white matter fiber loss, hypercellularity, and inflammation. While axonal injury was present in the CST and unmyelinated CC, myelin injury occurred only in the CST. The OPRA predominantly showed inflammation with myelin preservation. White matter cellular infiltration occurred in all tracts. Postmortem immunohistochemistry confirmed the imaging findings of decreased axonal fiber density, sparser fiber architecture, and increased cellular infiltration. Larger ventricular size was associated with greater white matter disruption. Building upon these results, DBSI provides an innovative approach for investigating the complex neuropathological effects of PHH on periventricular white matter microstructure.

## INTRODUCTION

Nearly 20% of infants who are born very preterm (≤32 weeks’ gestation) sustain spontaneous intraventricular hemorrhage (IVH)^1-3^. High-grade IVH occurs in up to one-half of these cases, with blood filling more than 50% of the ventricular volume with or without intraparenchymal extension^4^. This results in severe disruption of cerebrospinal fluid (CSF) dynamics, leading to CSF accumulation, raised intracranial pressure, and post-hemorrhagic hydrocephalus (PHH)^5^. Critically, PHH is associated with cognitive deficits in >85% of affected infants, cerebral palsy in 70%, and is a leading cause of epilepsy and hearing, visual, and speech impairments^1, 6^.

Many of the neurological deficits in PHH infants are attributed to injury of critical periventricular white matter (PVWM) structures, including the corpus callosum (CC), corticospinal tracts (CST), and optic radiations (OPRA)^7, 8^. Diffusion tensor imaging (DTI) has been widely used to demonstrate PVWM injury in PHH and across forms of neonatal hydrocephalus^8, 9^. DTI findings in PHH include reduced fractional anisotropy suggesting white matter disruption, reduced axial diffusivity reflecting axonal injury, and increased radial diffusivity reflecting myelin injury. These abnormal DTI measures in PHH correlate with poor neurodevelopmental outcomes in affected infants^8^. Nevertheless, DTI has inherent limitations in its ability to delineate complex white matter pathologies with adequate sensitivity and specificity^10, 11^. Critically, DTI is not capable of resolving diffusivities of crossing fibers^12, 13^, quantifying axonal density, or accurately assessing myelin integrity in the mixed presence of myelinated, dysmyelinated, and unmyelinated axons^13, 14^. Further, neuroinflammation, a hallmark of PHH pathophysiology, is associated with multiple extra-fiber water compartments, varying cell densities, edema, and axonal loss. Importantly, DTI’s inability to accurately differentiate these pathologies limit its efficacy for investigating PHH-related injury^15^.

Diffusion basis spectrum imaging (DBSI), a multi-tensor diffusion MRI (dMRI) analysis approach, addresses these key DTI limitations. In DBSI, water displacements are represented as multiple diffusion tensors reflecting different water compartments. This allows for separation of crossing fibers and provides measures of crossing angles, directional diffusivity of individual crossing fibers, and axonal loss^16^. Further, in addition to anisotropic diffusion compartments, the DBSI model includes isotropic tensors reflecting water compartments accounting for inflammatory cells and edema^17, 18^. DBSI has been validated in preclinical models of white matter injury^19, 20^ and utilized in adults to investigate clinical populations with varied white matter pathologies^16, 21^.

Leveraging DBSI’s unique strengths, we evaluated white matter integrity in a prospectively acquired, case-control cohort of very preterm infants with and without high-grade IVH and/or PHH. The aims of the study were to: 1) compare DBSI measures in the CC, CST, and OPRA of PHH infants to very preterm infants with and without IVH; 2) associate those DBSI measures with ventricular size and determine if the associations differed by group; and 3) compare postmortem histology between PHH and very preterm infants with and without IVH to confirm the MRI findings. We hypothesized that, compared to controls, the PVWM in PHH infants has increased cellularity, reduced fiber density, and disrupted fiber integrity, as confirmed by DBSI and histology, and larger ventricular size is associated with greater magnitude of PVWM disruption.

## METHODS

### Subjects

Very preterm infants who were screened for IVH via serial head ultrasounds obtained on day of life 3 and 7-10 as part of routine clinical care were prospectively recruited between 2007 and 2016 from a Level IV Neonatal Intensive Care Unit (NICU). Based upon head ultrasound findings and clinical course, recruited infants were categorized into three groups: VPT, IVH, and PHH. The VPT group had no identifiable brain injury or low-grade (Papile grade I or II) IVH^22^ on cranial ultrasound. The IVH group was comprised of VPT infants who sustained high-grade (Papile grade III or IV) IVH^22^, but did not develop hydrocephalus. Head ultrasounds were at minimum repeated bi-weekly for 21 days following IVH diagnosis as part of routine clinical care to monitor for the development of PHH.

The Hydrocephalus Clinical Research Network (HCRN) consensus criteria^23^ were used to identify the PHH group, which included IVH infants who developed hydrocephalus which required surgical treatment. Inclusion criteria for the PHH group were high-grade IVH; >72-hour life expectancy; frontal-occipital horn ratio (FOHR) ≥.55; and two of the following: bulging fontanel (above the level of surrounding bone), split sagittal sutures (≥2 mm in mid-parietal region) or ≥3 episodes of documented bradycardia over 24 hours. Treatment of the PHH infants involved two distinct phases: a temporizing phase to permit ventricular decompression prior to term equivalent postmenstrual age and a permanent phase to enable long-term CSF diversion with a ventriculoperitoneal shunt or endoscopic third ventriculostomy (ETV). The decision to perform a permanent CSF diversionary procedure was made using the following HCRN consensus criteria: weight ≥1800 g; abdomen favorable (no drains or necrotizing enterocolitis); and persistent CSF removal required to maintain a) the anterior fontanel at/below the level of surrounding bone, b) splaying of sagittal suture <2 mm, and c) FOHR ≤ .55. Choroid plexus cauterization (CPC) was not performed because the infants in this cohort who underwent ETV were treated prior to CPC’s adaptation as common practice at our institution.

Informed written consent was provided by parents prior to each subject’s enrollment in the study. All protocols and procedures were approved in advance by the Washington University Human Research Protection Office.

### Perinatal clinical factors

To minimize confounding factors, all infants who had congenital infections (e.g., cytomegalovirus, toxoplasma, rubella) or chromosomal abnormalities were excluded. In addition, assessment of clinical health status was standardly assessed across subjects using a well-established clinical risk index score^24^ that assigned a 1 or 0 for the presence or absence of discrete clinical factors, respectively. The ten factors assessed were: 1) small for gestational age at birth or intrauterine growth restriction; 2) oxygen therapy at 36 weeks; 3) postnatal steroids; 4) necrotizing enterocolitis; 5) patent ductus arteriosus requiring indomethacin or ibuprofen therapy; 6) retinopathy of prematurity of any grade or stage; 7) culture-positive neonatal sepsis; 8) weight-for-length ratio at term equivalent MRI acquisition >3 standard deviations below the measured ratio at birth; 9) duration of receiving total parenteral nutrition >75^th^ percentile and 10) mother did not receive antenatal steroids. Scores from the 10 factors were added to obtain a composite risk index score^24^.

### Image acquisition

All MRI studies were acquired without sedation following institutional guidelines^25^. All infants were scanned at/near term equivalent (35-43 weeks’ postmenstrual age) on a 3T Siemens Trio MRI system (Erlangen, Germany), using an infant-specific, quadrature head coil (Advanced Imaging Research, Cleveland, OH, USA). Anatomical images including T1W (repetition time [TR] 1550 ms, echo time [TE] 3.05 ms, 1.0×1.0×1.3 mm^3^ spatial resolution) and T2W (TR 8210 ms, TE 161 ms, 1.0×1.0×1.0 mm^3^ spatial resolution) data were obtained. dMRI data (TR 13300 ms, TE 112 ms, 1266 Hz/Px bandwidth, 128 mm field of view, 1.2×1.2×1.2 mm^3^ spatial resolution) with 27-48 b-directions and amplitudes ranging 0-1200 s/mm^2^ were also collected.

### Image processing

The diffusion signal attenuation curve was modeled as a mono-exponential function plus a constant, and diffusion parameters were estimated using Bayesian Probability theory^26^. dMRI data were processed using FSL tools^27^ and corrected for eddy current distortions using Eddy^28^, susceptibility induced distortions using ApplyTopup^28^, and motion corrected using FLIRT^29^. The CC, bilateral posterior limbs of the internal capsules, designated CST, and bilateral OPRA were selected *a priori* as our regions of interest (ROIs) as they have been shown to demonstrate improvement in dMRI indices following hydrocephalus treatment and relate to PHH-related neurodevelopmental outcomes^8, 30^.

For each subject, linear ventricular size was measured by two independent raters on axial T2W images using the FOHR approach^31^. Reproducibility of these measures had been previously validated^32^. Each ROI was segmented on three contiguous axial slices to minimize through-slice partial volume averaging using Analyze 10.0 software (AnalyzeDirect, Inc., Overland Park, KS, USA) (Fig. 1).

**Fig. 1.**
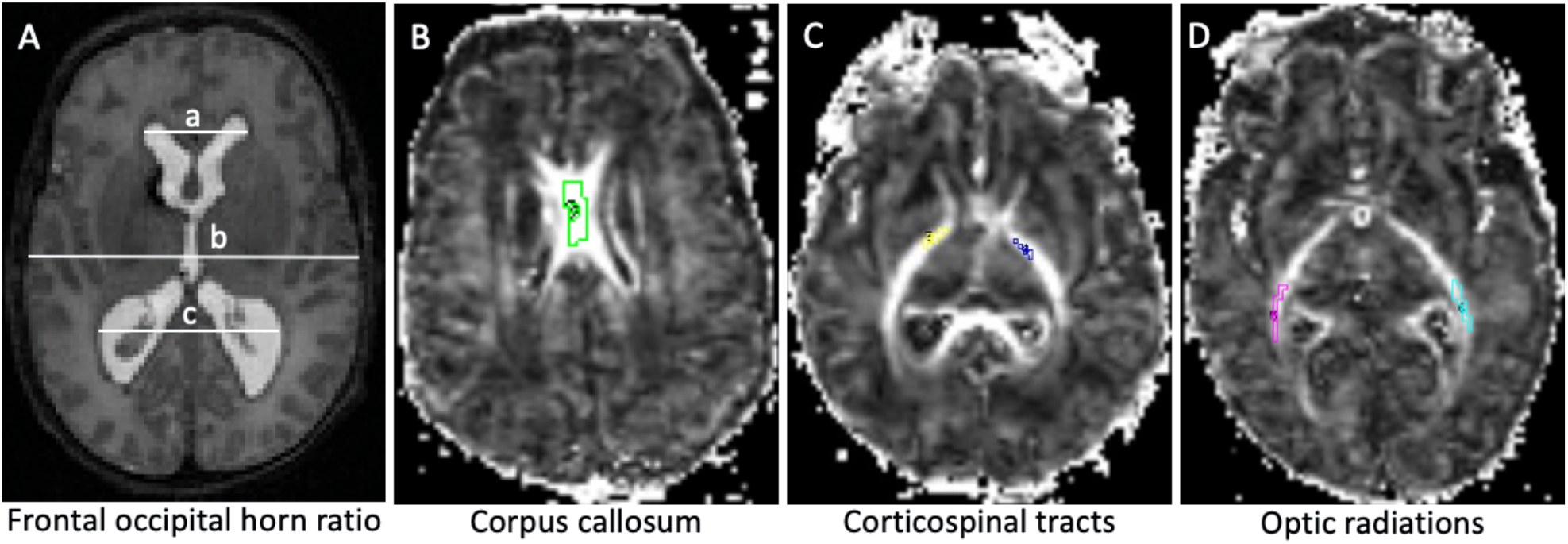
Representative axial T2W MRI scan (A) and contiguous axial fractional anisotropy (FA) maps (B-D) of preterm infants. Ventricular size was determined by the Frontal-Occipital Horn Ratio which measures ventricular size as the sum of the distance between the widest lateral walls of the frontal and occipital horns (a + c), divided by twice the widest biparietal diameter at the level of the foramen of Monro (b). On the FA maps, segments of the white matter bundles of interest within the corpus callosum, bilateral corticospinal tracts and bilateral optic radiations are demarcated.

The preprocessed 27-48 directional dMRI images were analyzed via the DBSI pipeline^17^ in MatLab® (MathWorks, Natick, MA, USA). As demonstrated in Equation 1, the DBSI model is a sum of the multiple anisotropic diffusion tensors (first term) and a spectrum of isotropic diffusion tensors (second term):

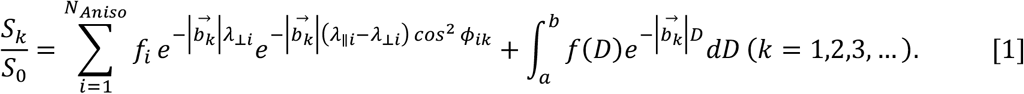

Here *b*_*k*_ is the *k*^th^ diffusion gradient; *S*_*k*_*/S*_*0*_ is the acquired diffusion weighted signal at direction of *b*_*k*_ normalized to non-diffusion-weighted signal; *N*_*Aniso*_ is the number of anisotropic tensors to be determined; *ϕ*_*ik*_ is the angle between the diffusion gradient *b*_*k*_ and the principal direction of the *i*^*th*^ anisotropic tensor; 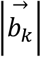 is *b*-value of the *k*^*th*^ diffusion gradient; *λ*_‖*i*_ and *λ*_*⊥i*_ are the axial and radial diffusivity of the *i*^*th*^ anisotropic tensor under the assumption of cylindrical symmetry; *f*_*i*_ is the signal-intensity-fraction (non-diffusion-weighted) of the *i*^*th*^ anisotropic tensor; *a, b* are low and high diffusivity limits of the isotropic diffusion spectrum; *f(D)* is the spectrum of signal-intensity-fraction (non-diffusion-weighted) at all possible isotropic diffusivity *D*.

*A priori* division of isotropic components was established using an apparent diffusion coefficient (ADC) cutoff of ≤.3 µm^2^/ms representing restricted diffusion, ADC values between .3-2.5 µm^2^/ms representing hindered diffusion, and ADC ≥2.5 µm^2^/ms representing free diffusion^16-21^. Following DBSI analysis, ROI-specific metrics were obtained via voxel-based computations. DBSI metrics included: fiber specific FA (FFA), AD (FAD), and RD (FRD); axonal density (fiber fraction, FF); non-fiber related restricted fraction (RF); and non-restricted fraction (NRF).

### Immunohistochemistry

Neurocytology was performed on 10 formalin-fixed post-mortem brain specimens: five VPT, three IVH, and two PHH (medical records had documentation supporting a PHH diagnosis prior to death based on the HCRN criteria^23^). The brains of two full-term infants who died of non-neurologic causes were similarly processed for comparison. None of these infants were part of the imaging arm of this study. Paraffin-embedded 5-µm sections of all patients containing the CC were stained with hematoxylin and eosin (H&E) to assess cellular structure, edema and cellularity. In addition, immunostaining for synaptophysin and GFAP (1:300, catalog #7260, Abcam) was performed to better assess neuropil and astrocytes respectively. To examine associations between PVWM and the subjacent ventricular (VZ) and subventricular (SVZ) zones, the VZ/SVZ regions from consecutive sections were additionally stained with H&E to evaluate the cytoarchitecture and immunostained with DAPI (D1306 Thermo Fisher Scientific) and GFAP using double-labeling to examine the immature ependymal cells and reactive astrocytes, and with DAPI and IBA-1 (1:100, #PA5-18039, Invitrogen) as a surrogate for microglia- and macrophage-associated elements. All slides were scanned using a Nanozoomer-XR (Hamamatsu Photonics). Qualitative comparisons were performed by a neuropathologist (SD). Percent VZ disruption was quantified as the cumulative length of the disrupted ependymal regions with the denominator being the entire length of VZ available on the same slide.

### Statistical Analysis

All statistical analyses were performed with *SAS version 9*.*4*. Numeric values were tested for normality using the Shapiro-Wilk test and box plots for visual examination, which suggested the use of parametric and non-parametric tests when applicable. Voxel-based DBSI measures in the right and left CST and OPRA were combined. Multiple comparisons with Kruskal-Wallis and Wilcoxon Rank-Sum tests with Bonferroni adjustments were performed between VPT, IVH, and PHH groups for all DBSI measures. To ensure appropriate comparability across groups, a generalized linear regression model was used to compare group differences while adjusting for the following covariates: 1) birth gestational age, 2) postmenstrual age at MRI scan, 3) number of diffusion-encoding directions, 4) clinical risk composite index, and 5) sex. The association of FOHR with DBSI variables was described by Pearson’s correlation coefficients. Bonferroni-corrected *p* values <.05 were considered significant.

## RESULTS

### Subjects

A total of 130 infants (VPT=91, IVH=20, PHH=19) with dMRI data were screened for possible inclusion in the study. Reasons for exclusion across groups included: positive maternal urine drug screen (VPT=1, IVH=1) and atypical pattern of brain injury/abnormal MRI finding (VPT=2). DBSI fitting could not be performed in two severely injured patients whose ROIs could not be accurately delineated. In addition, seven infants were excluded because they had severe white matter cysts that caused significant changes in ventricular morphology near the foramen of Monro precluding accurate ventricular size measurements. The scans of seven infants could not be fitted with DBSI due to having less than 25 diffusion-encoding directions and/or b-values. Among 95 infants (48 males, 47 females) included in the final analyses, there were 68 VPT, 15 IVH, and 12 PHH infants (Fig. 2).

**Fig. 2.**
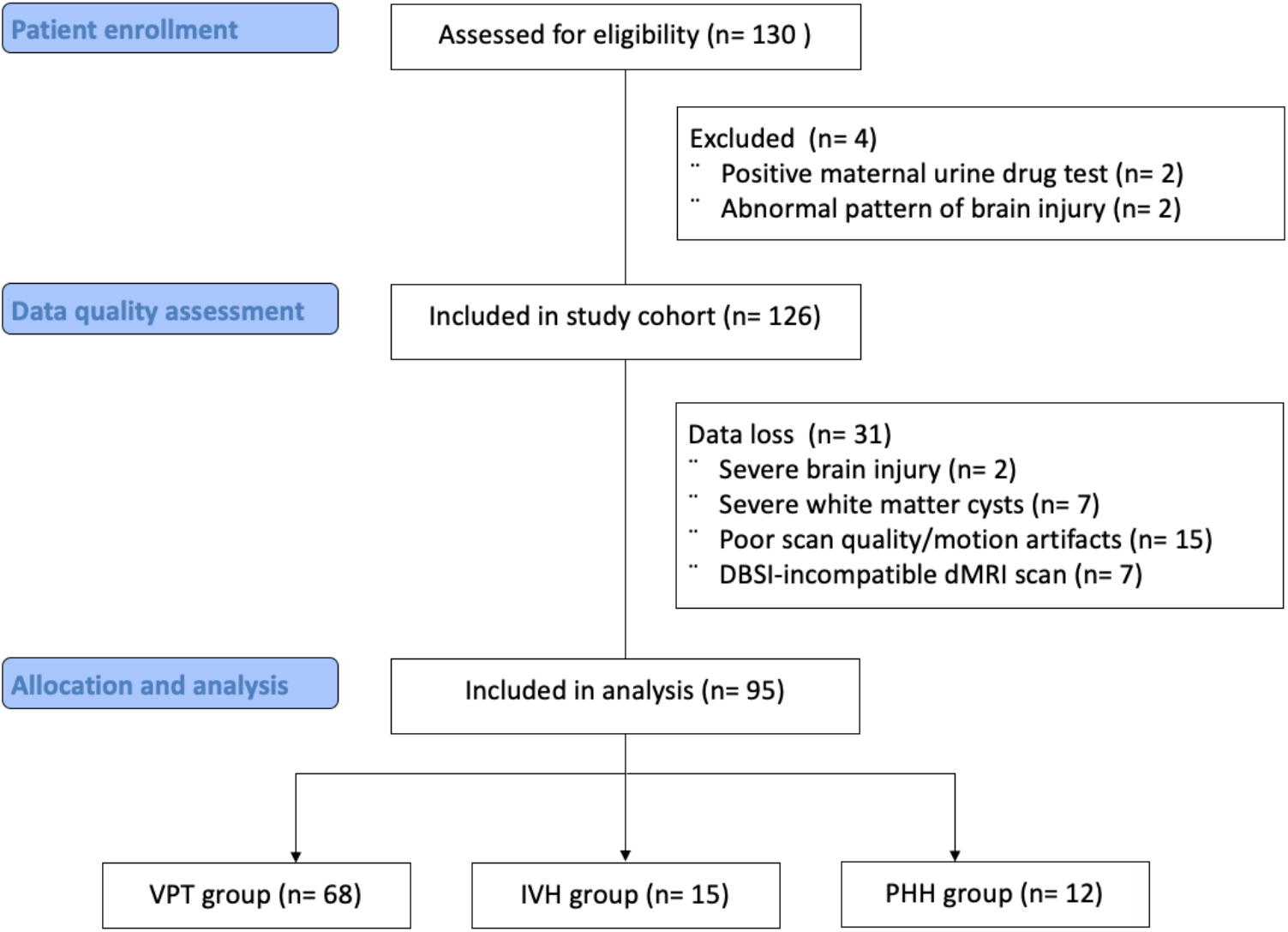
Flowchart of patient enrolment and selection for study inclusion. *VPT = very preterm; IVH = intraventricular hemorrhage; PHH = posthemorrhagic hydrocephalus; DBSI= diffusion basis spectrum imaging; dMRI = diffusion MRI*

The median gestational age at birth and postmenstrual age at dMRI acquisition across groups were 27±2 and 38±2 weeks, respectively. There were no between-group differences in median birth weight (910±390 g, p=.350). Median of the composite clinical risk scores were similar among IVH (3, IQR = 2) and PHH (3, IQR = 4) infants, whereas that those for the VPT infants were lower (1, IQR = 3) (Table 1). Prior to MRI, all 12 PHH infants had undergone temporary ventricular access device placement, including 10 of which had undergone permanent VP shunt placement and two of which had undergone ETV to treat their hydrocephalus. By convention, the PHH infants had larger ventricles than the VPT and IVH groups (median FOHR .52±.10 versus .41±.03) (Fig. 3). On subgroup analysis, there were no differences between the shunt- and ETV-treated infants on clinical characteristics and DBSI measures.

**Table 1.** Baseline characteristics of 95 preterm infants

|  | VPT | IVH | PHH |
| --- | --- | --- | --- |
| # of patients | 68 | 15 | 12 |
| Sex |  |  |  |
| # of male infants | 29 | 7 | 12 |
| # of female infants | 39 | 8 | 0 |
| GA at birth (weeks) | $27.0\pm 2.8$ | $25.0\pm 3.0$ | $24.5\pm 4.0$ |
| PMA at dMRI acquisition (weeks) | $37.0\pm 1.0$ | $38.0\pm 4.0$ | $39.0\pm 1.0$ |
| Birth weight (g) | $932.5\pm 350.0$ | $740.0\pm 330.0$ | $807.5\pm 304.5$ |
| Frontal-occipital horn ratio | $.41\pm .03$ | $.41\pm .04$ | $.52\pm .10$ |
| Clinical risk score | $1\pm 3$ | $3\pm 2$ | $3\pm 4$ |

*All continuous variables are reported as median ± interquartile range within the respective groups. VPT = very preterm; IVH = intraventricular hemorrhage; PHH = post-hemorrhagic hydrocephalus; GA = gestational age; PMA = postmenstrual age*

**Fig 3.**
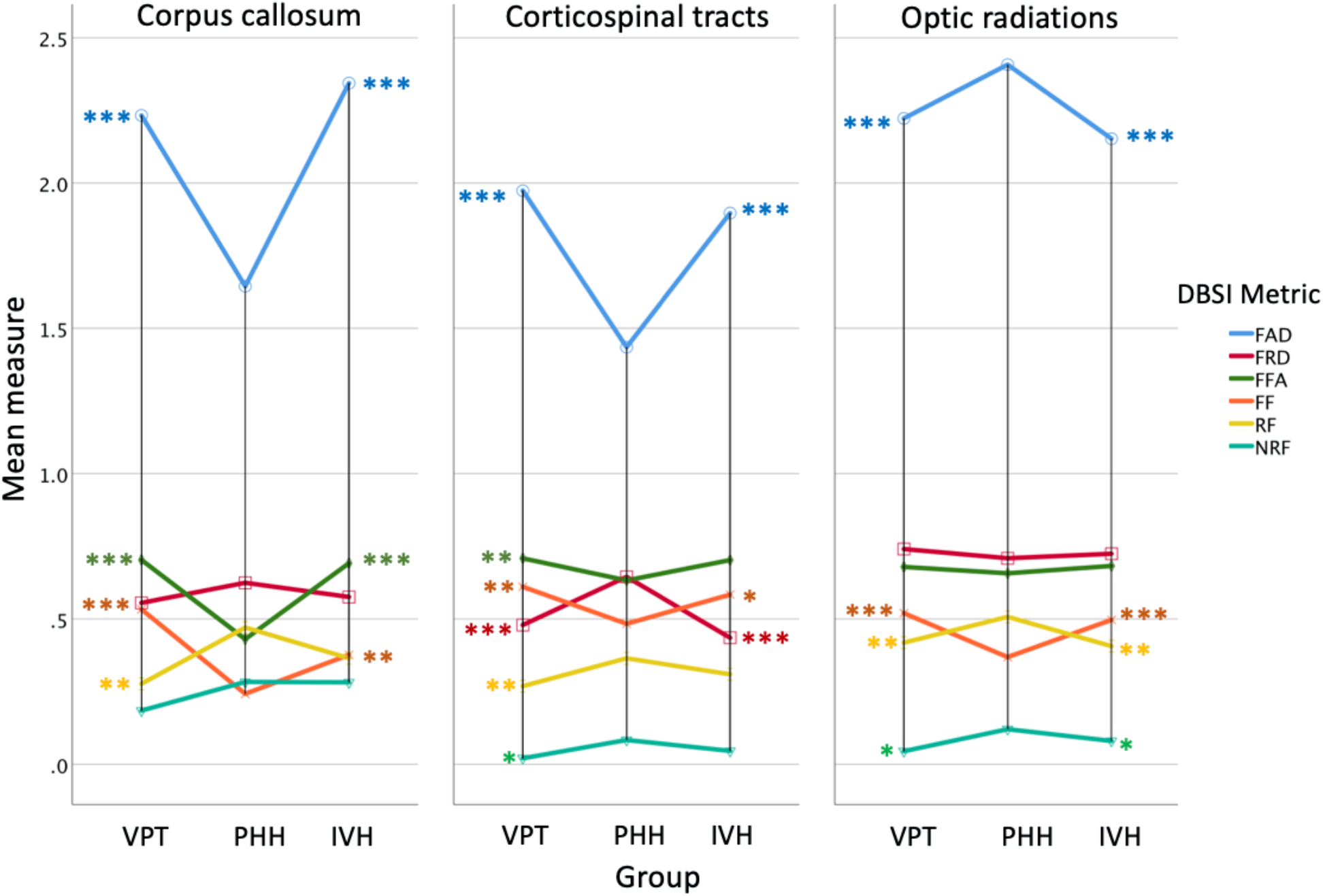
DBSI measures between very preterm infants without brain injury (VPT), infants with high-grade intraventricular hemorrhage (IVH), and infants with posthemorrhagic hydrocephalus (PHH) requiring treatment in three critical white matter tracts. While the tract-specific patterns were variable, in comparison to VPT and IVH, PHH infants had the most severe white matter disruption marked by the ***lowest*** fiber fraction (FF; measure fiber density), fiber fractional anisotropy (FFA; measure of the directional dependence of water diffusivity), and fiber axial diffusivity (FAD; measure of the rate of water diffusion parallel to axons), as well as the ***highest*** measures of fiber radial diffusivity (FRD; measure of the rate of water diffusion perpendicular to axons), and markers of inflammation reflected as restricted fraction (RF; measure of cellular infiltration) and non-restricted fractions (NRF; measure of vasogenic edema). All comparisons are Bonferroni corrected with *\*\*\*, ** and * representing p-values <*.*001, <*.*01, and <*.*05, respectively, between PHH and the corresponding group*.

### Corpus callosum (CC)

In the CC, PHH had the lowest FAD (*p*<.001) and there was no difference between IVH and VPT. There were no between-group differences for FRD. PHH had the lowest FFA (*p*<.001) and there was no difference between IVH and VPT. The fiber densities in the PHH group were 55% (*p*<.001) and 37% (*p*=.004) lower than the VPT and IVH groups, respectively. The IVH group had 28% lower FF than the VPT group (*p*=.021). While the PHH group had higher RF than VPT group (*p*=.002), there was no difference either between PHH and IVH or IVH and VPT. There were no between-group differences in NRF (Fig. 3 and Table 2).

**Table 2.**
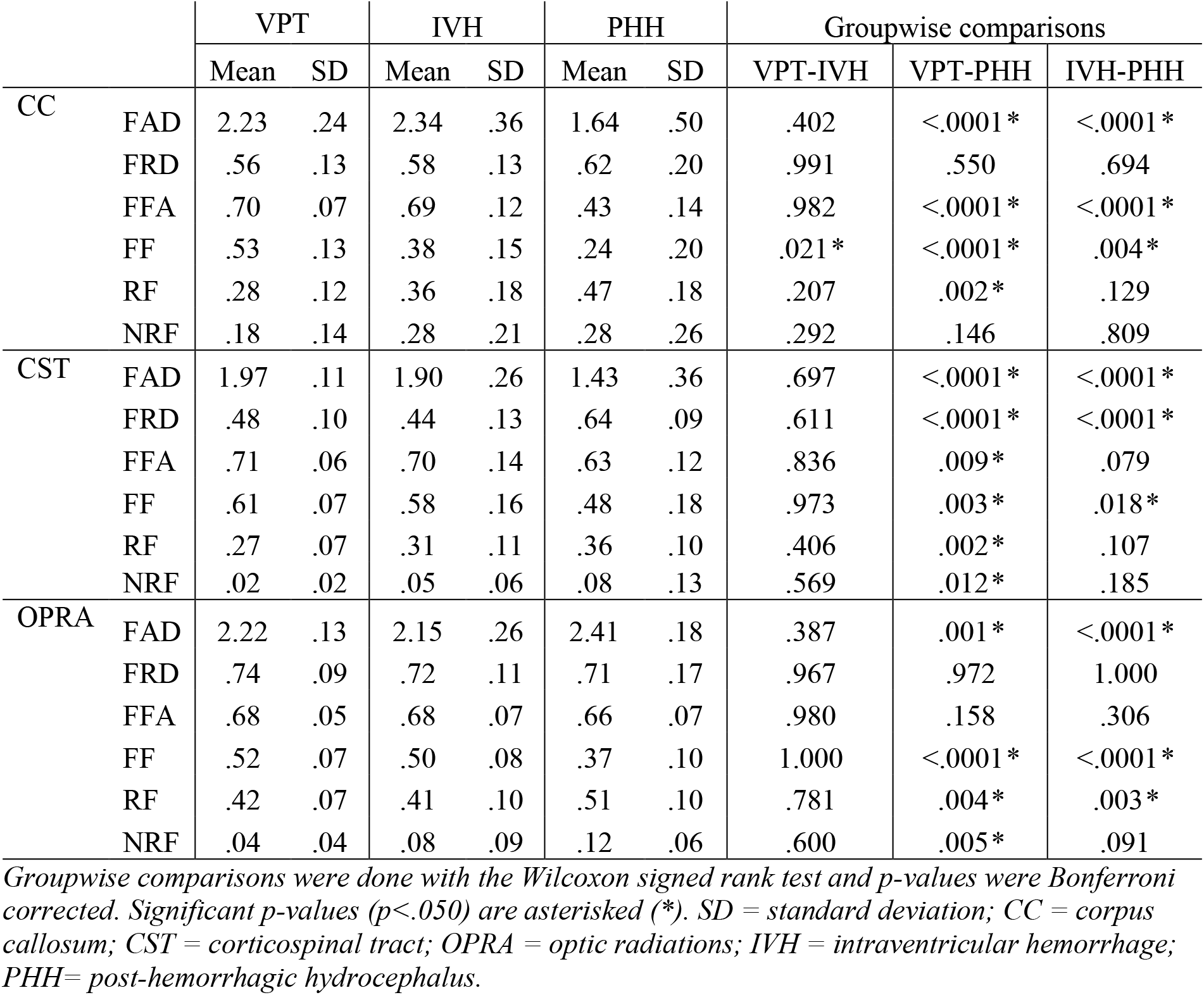
DBSI measures in very preterm infants with and without IVH/PHH

|  |  | VPT |  | IVH |  | PHH |  | Groupwise comparisons |  |  |
| --- | --- | --- | --- | --- | --- | --- | --- | --- | --- | --- |
|  |  | Mean | SD | Mean | SD | Mean | SD | VPT-IVH | VPT-PHH | IVH-PHH |
| CC | FAD | 2.23 | .24 | 2.34 | .36 | 1.64 | .50 | .402 | <.0001* | <.0001* |
|  | FRD | .56 | .13 | .58 | .13 | .62 | .20 | .991 | .550 | .694 |
|  | FFA | .70 | .07 | .69 | .12 | .43 | .14 | .982 | <.0001* | <.0001* |
|  | FF | .53 | .13 | .38 | .15 | .24 | .20 | .021* | <.0001* | .004* |
|  | RF | .28 | .12 | .36 | .18 | .47 | .18 | .207 | .002* | .129 |
|  | NRF | .18 | .14 | .28 | .21 | .28 | .26 | .292 | .146 | .809 |
| CST | FAD | 1.97 | .11 | 1.90 | .26 | 1.43 | .36 | .697 | <.0001* | <.0001* |
|  | FRD | .48 | .10 | .44 | .13 | .64 | .09 | .611 | <.0001* | <.0001* |
|  | FFA | .71 | .06 | .70 | .14 | .63 | .12 | .836 | .009* | .079 |
|  | FF | .61 | .07 | .58 | .16 | .48 | .18 | .973 | .003* | .018* |
|  | RF | .27 | .07 | .31 | .11 | .36 | .10 | .406 | .002* | .107 |
|  | NRF | .02 | .02 | .05 | .06 | .08 | .13 | .569 | .012* | .185 |
| OPRA | FAD | 2.22 | .13 | 2.15 | .26 | 2.41 | .18 | .387 | .001* | <.0001* |
|  | FRD | .74 | .09 | .72 | .11 | .71 | .17 | .967 | .972 | 1.000 |
|  | FFA | .68 | .05 | .68 | .07 | .66 | .07 | .980 | .158 | .306 |
|  | FF | .52 | .07 | .50 | .08 | .37 | .10 | 1.000 | <.0001* | <.0001* |
|  | RF | .42 | .07 | .41 | .10 | .51 | .10 | .781 | .004* | .003* |
|  | NRF | .04 | .04 | .08 | .09 | .12 | .06 | .600 | .005* | .091 |

Groupwise comparisons were done with the Wilcoxon signed rank test and *p*-values were Bonferroni corrected. Significant *p*-values ( $p < .050$ ) are asterisked (\*). SD = standard deviation; CC = corpus callosum; CST = corticospinal tract; OPRA = optic radiations; IVH = intraventricular hemorrhage; PHH = post-hemorrhagic hydrocephalus.

### Corticospinal tracts (CST)

In the CST, fiber-specific metrics demonstrated PHH had the lowest FAD (*p*<.001) and highest FRD (*p*<.001), with no difference between IVH and VPT across both measures. Although the PHH group had lower FFA than VPT group (*p*=.010), there was no difference either between PHH and IVH (p=.079) or IVH and VPT (p=.836) groups. Fiber density analyses showed that the PHH group had 21% (*p*=.003) and 17% (*p*=.018) less FF than VPT and IVH, respectively. There was no difference in fiber density between IVH and VPT. Isotropic spectrum analyses demonstrated that the PHH group had higher RF (p=.002) and NRF (.012) than the VPT group; however, there was no difference either between PHH and IVH or between IVH and VPT on both measures (Fig. 3 and Table 2).

### Optic radiations (OPRA)

In the OPRA, fiber level analysis showed that PHH had the highest FAD (*p*<.001) with no difference between IVH and VPT. No between group differences were found for FRD and FFA. Fiber density analysis showed that PHH had 29% lower FF than VPT (*p*<.001) and 26% lower than the IVH (*p*<.001), with no difference between the IVH and VPT groups. Isotropic analyses showed that PHH had the highest RF and NRF. However, there was no difference between IVH and VPT on RF measures, as well as no difference between either PHH and IVH (*p*=.091) nor VT and IVH (*p*=.600) (Fig. 3 and Table 2).

### Ventricular size-DBSI Correlations

Across all three white matter tracts, greater ventricular size was correlated with greater magnitude of abnormality in DBSI measures with variable patterns. In the CC, FOHR related negatively with FAD (*p*<.001), FFA (*p*<.001), and FF (*p*<.001) and positively with RF (*p*<.001). There was no correlation between FOHR and FRD or NRF in the CC. Similar to the CC, FOHR related negatively to both FAD (*p*<.001) and FFA (*p*<.001). Positive correlations were observed between FOHR and NRF (p=.011) and FRD (p<.001) in the CST. There were no correlations between FOHR and FF or RF in the CST. In the OPRA, FOHR related positively with FAD (*p*<.001) and NRF (*p*=.006) and negatively with FF (*p*=.031). No correlations were found between FOHR and FRD, FFA, or RF in the OPRA (Fig. 4).

**Fig 4.**
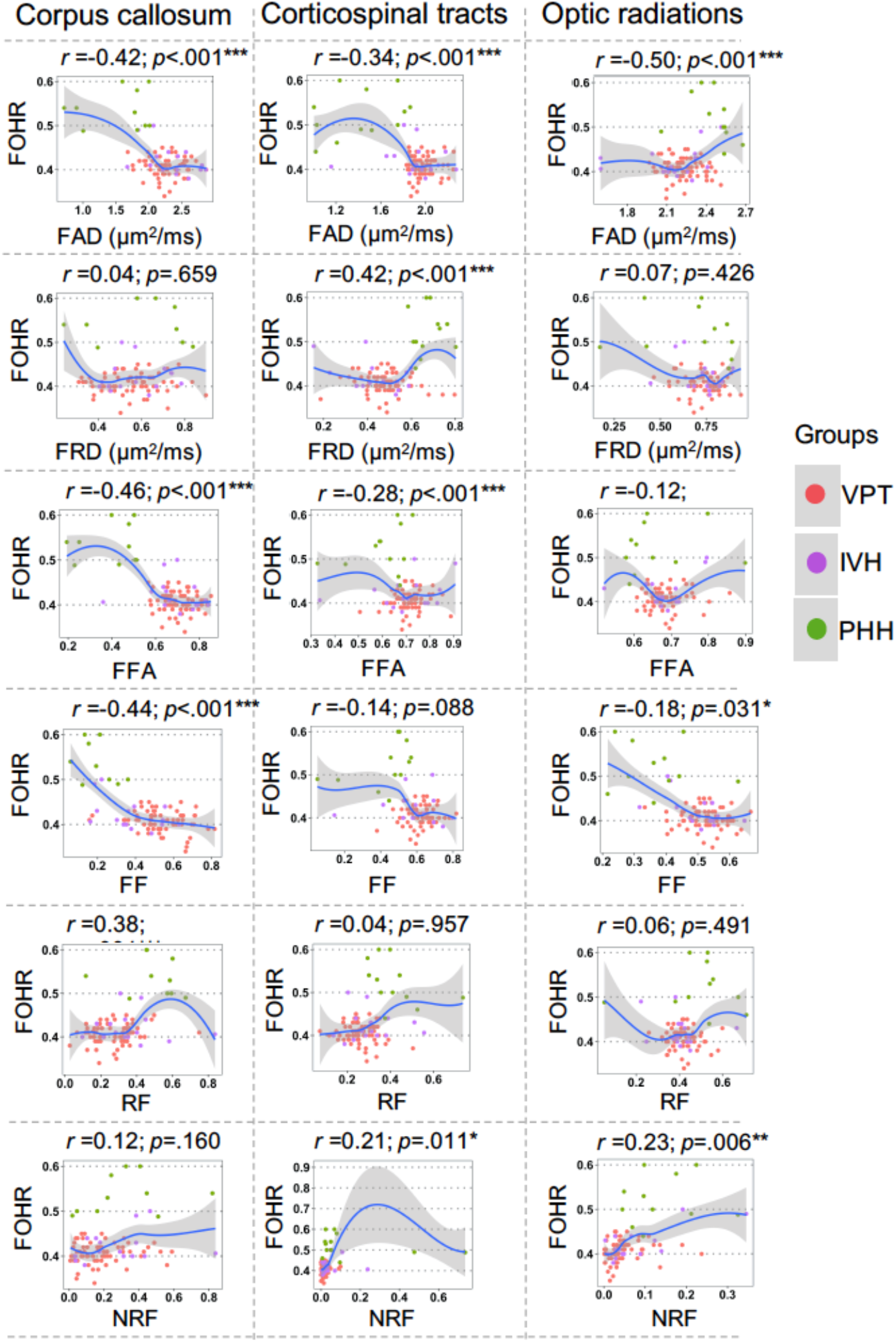
Ventricular size (frontal-occipital horn ratio, FOHR<u>^43^</u>) and dMRI correlations between very preterm infants without brain injury (VPT), infants with high-grade intraventricular hemorrhage (IVH), and infants with posthemorrhagic hydrocephalus (PHH) requiring treatment in three critical white matter tracts. While the tract-specific patterns were variable, increase in ventricular size was associated with greater abnormalities in dMRI measures, reflected as positive correlations between FOHR and restricted fraction (RF; measure of cellular infiltration), non-restricted fractions (NRF; measure of vasogenic edema), and fiber radial diffusivity (FRD; measure of the rate of water diffusion perpendicular to axons), as well as negative correlations with fiber fractional anisotropy (FFA, measure of the directional dependence of water diffusivity), fiber axial diffusivity (FAD; measure of the rate of water diffusion parallel to axons), and fiber fraction (FF; measure fiber density). All comparisons are Bonferroni corrected with *\*\*\*, **, and * representing p-values <*.*001, <*.*01, and <*.*05, respectively*.

### Postmortem tissue analyses

In comparison to VPT and IVH, PHH infants demonstrated white matter tracts with higher cellularity. In addition, PHH infants had decreased synaptophysin staining and variable amounts of cytoplasmic vacuolation (Fig. 5).

**Fig. 5.**
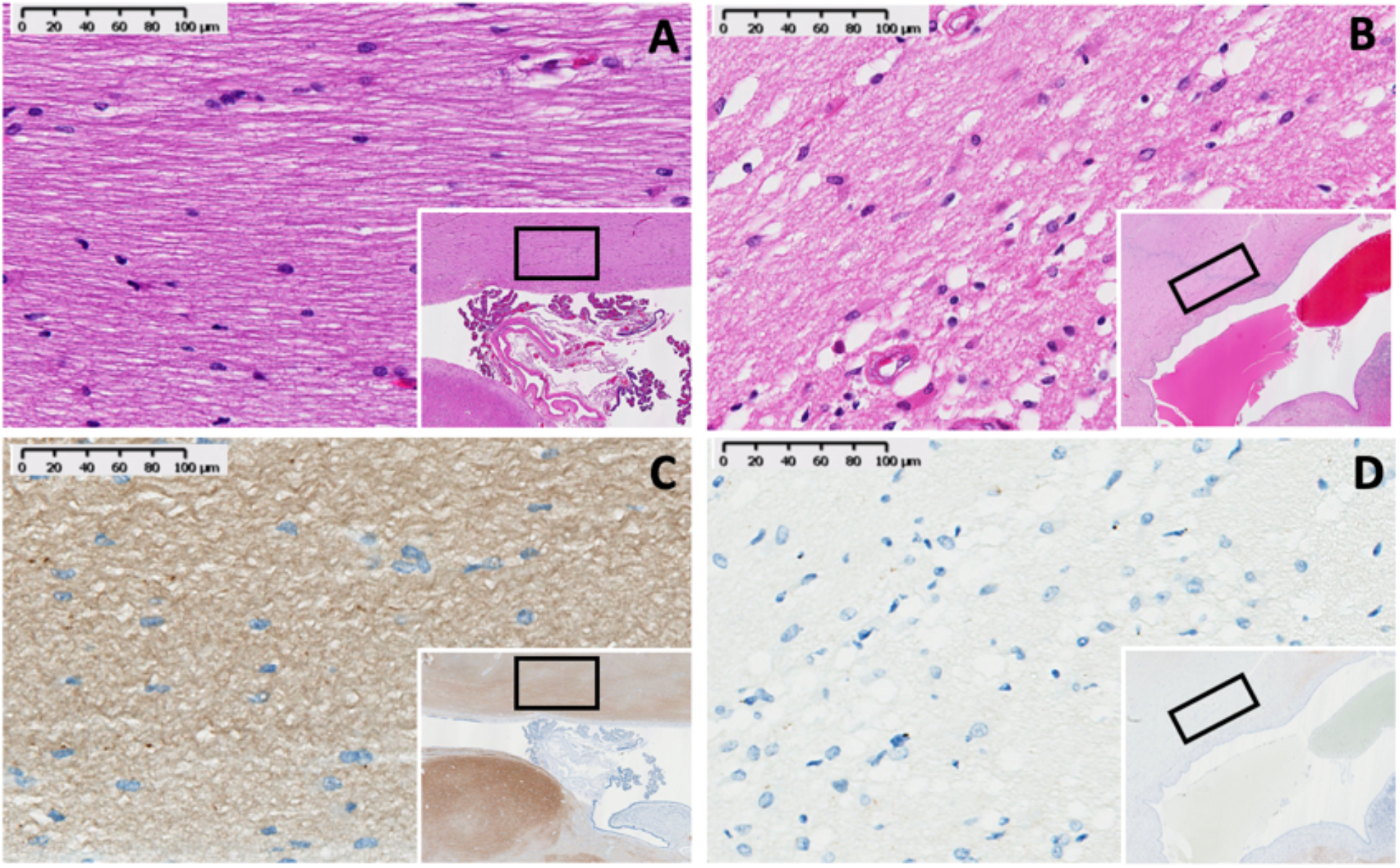
Representative histological staining of post-mortem tissue from corpus callosum (CC) of preterm infants. H&E (A, B) and synaptophysin (C, D) of preterm infants without (A, C) or with (B, D) PHH. PHH infants had relatively increased cellular infiltration (B, D) and vacuolation of their cytoplasm to suggest edema (B). Decreased axonal staining in PHH (D) suggests a relatively decreased fiber density in comparison to controls (C). The CC is not myelinated at term equivalent.

Analyses of the adjoining VZ/SVZ of IVH, VPT, PHH in comparison to that of full term infants who died from non-neurologic causes (FT) demonstrated that IVH, and to a greater extent PHH, were associated with VZ/SVZ disorganization on H&E staining and VZ/SVZ disruption when compared to VPT (*p*=.01), IVH (*p*=.16), and FT (*p*=.01) infants. Both IVH and PHH were associated with disruption of radial glial cells in the VZ, characterized by loss of their basal projections and VZ lattice. There was increased expression of GFAP and IBA1 in the VZ/SVZ regions of PHH when compared to the FT and VPT infants. The difference in percent disruption between IVH and VPT infants was not significant (*p*=.34). Compared to the IVH and PHH tissues, which had established VZ disruption with denuded ependyma and no cilia, regions of disruption in VPT tissues were predominantly at early stages and characterized by VZ protrusions toward the ventricular space with preserved cilia (Fig. 6).

**Fig 6.**
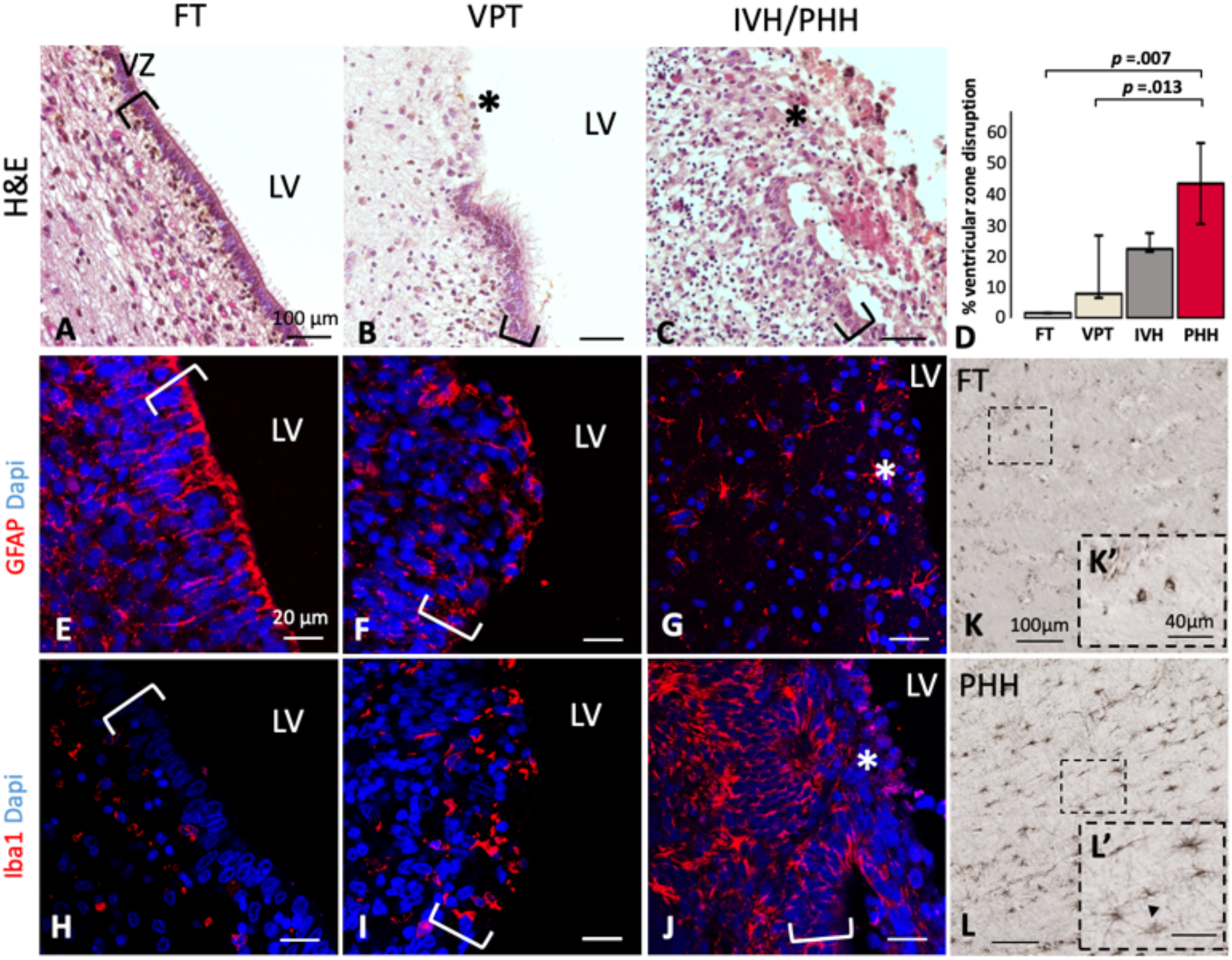
Representative immunohistochemistry of postmortem brain tissue from a full-term control (FT: A, E, H and K), very preterm with low-grade intraventricular hemorrhage (VPT; B, F and I), and very preterm with high-grade intraventricular hemorrhage (IVH/PHH: C, G, J, and L) infants. Asterisk (*) indicate regions of disruption. The H&E images (A, B, and C) demonstrate an intact ventricular zone (VZ) with normal multiciliated ependymal cells (A), a region undergoing early VZ disruption with protrusion of the VZ toward the lateral ventricular (LV) space with preserved cilia (B), and an established VZ disruption with denuded ependyma and no cilia (C). Percent VZ disruption, quantified as the cumulative length of the disrupted ependymal regions as a factor of the entire length of VZ available on the same slide, demonstrate PHH was associated with more severe VZ damage than IVH, VPT, and FT infants (D). Adjacent slides of the same infants double-labeled with anti-GFAP and DAPI demonstrate the characteristic shape of radial glial cells with basal projections (E) undergoing progressive changes in the pattern of GFAP expression, initially showing a lack of basal projections (F) and subsequently a proliferation of GFAP-positive cells demonstrating reactive astrocytosis (G). Anti-Iba1 and DAPI immunolabelling of adjacent slides (H, I, and J) demonstrate reclusion of microglia/macrophages, mainly in the subventricular zone (H) in the FT infant, but predominance of Iba1 positive cells, mostly in the area of VZ disruption (I), which was worse in IVH and PHH infants (J). Examination of the adjacent white matter region in the corpus callosum of FT (K) and PHH (L) infants demonstrated increased GFAP expression in the PHH infant. Morphologically, the GFAP-positive cells in the FT infant did not appear reactive (K’) when compared to the reactive astrocytosis observed in the PHH infant (L’, arrow). Asterisks indicate disrupted VZ regions. Scale bars A-J (20 µm), K & L (100 µm) and K’ & L’ (40 µm).

## DISCUSSION

### Tract-specific white matter injury associated with PHH

In comparison to IVH and VPT groups, infants with PHH demonstrated the most severe white matter abnormalities across DBSI measures. There were no consistent differences in measures between the IVH and VPT groups, a finding which indicates the deviations observed in the PHH group were associated with hydrocephalus rather than the antecedent hemorrhage or preterm birth. Key tract-specific white matter injury patterns in PHH included reduced fiber density in the setting of axonal and/or myelin injury, increased cellular infiltration, vasogenic edema, and inflammation. Loss of white matter fibers occurred across all three tracts in PHH infants, and all tracts demonstrated evidence of high cellular infiltration. Measures of axonal injury were highest in the unmyelinated CC. In the myelinated CST, both axonal and myelin injury were observed. In the OPRA, axonal and myelin integrity were preserved in the setting of increased extra-fiber cellular infiltration (RF) and edema (NRF), indicating there was increased extracellular space due to fiber loss and transependymal CSF migration^32^ (Fig. 7). Of note, IVH independently had a detrimental effect on fiber density in the CC, likely from the direct effects of blood breakdown products in the CSF and/or reflecting that more severe IVH is more likely to progress to PHH. Increasing ventricular size correlated with worse DBSI metrics, providing objective, quantifiable data that can be incorporated into long-standing discussions of the role of ventriculomegaly in PHH-related neurological disability^33^. DBSI findings of decreased fiber density, sparser architecture, increased cellular infiltration, edema, and inflammation in periventricular white matter tracts were confirmed on immunohistochemistry in postmortem specimens.

**Fig. 7.**
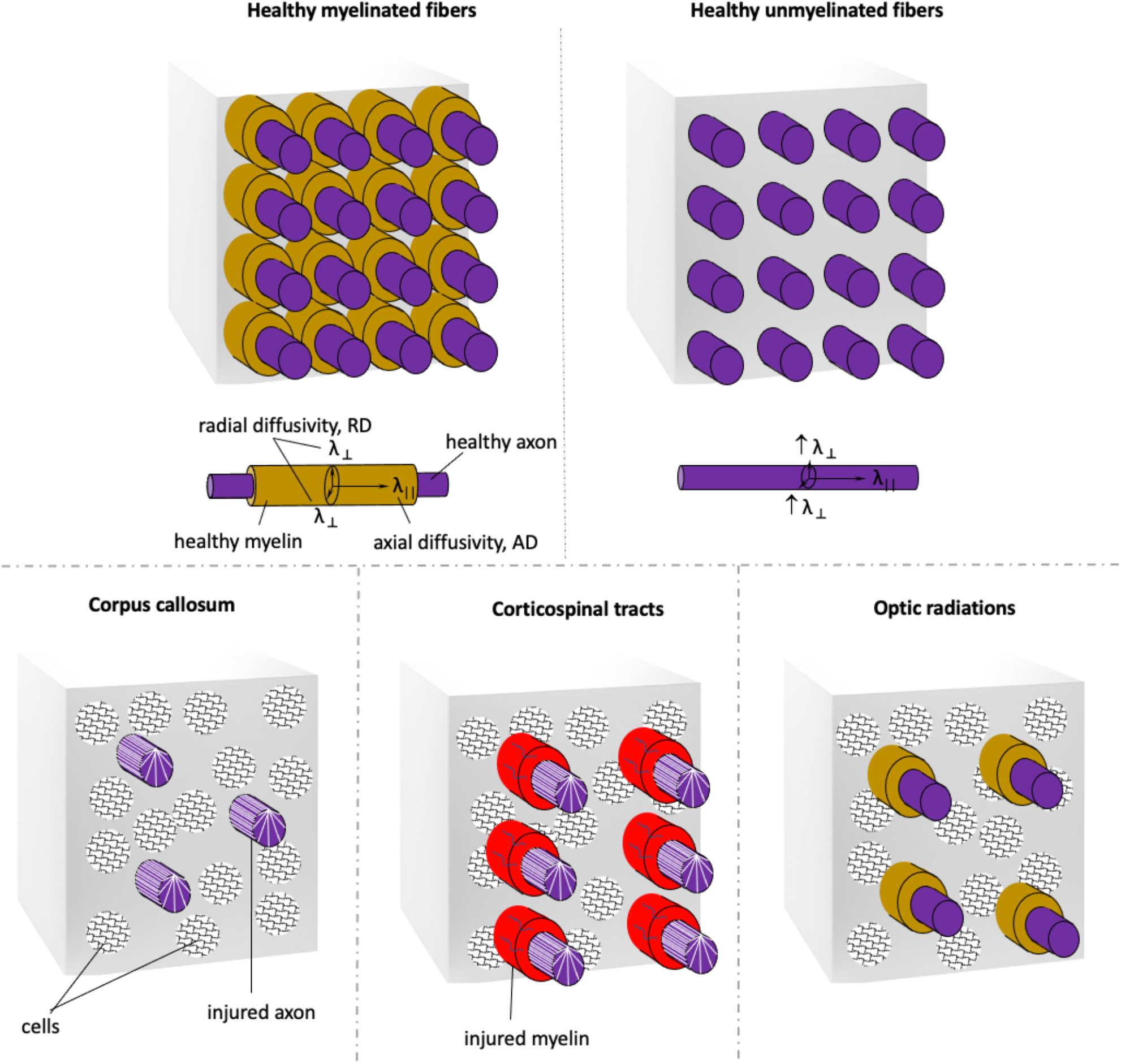
A schematic rendering of postulated pathology underlying the white matter bundles detected with dMRI in the three tracts is shown. The overall MRI voxels from healthy myelinated and unmyelinated white matter are shown in the top row. The bottom row shows voxels from the corpus callosum, corticospinal tracts and optic radiations of PHH infants, drawn to same scale as the healthy voxels, to demonstrate relative fiber density, inflammatory cells, and estimated inter-axonal spacing. The corpus callosum is not myelinated at this developmental stage. Relative to healthy white matter, there was significantly reduced fiber density in all tracts, which was worst in the corpus callosum. The striated axons of callosum, corticospinal tracts indicate axonal injury. In the corticospinal tract, the increased radial diffusivity suggests myelin injury, which is represented as red sleeves around the axons. In the isotropic spectrum, there was increased cellularity in the corpus callosum, corticospinal tract and optic radiations.

### Effect of PHH on early brain and white matter development

While this study is the first to utilize DBSI to investigate white matter microstructure in pediatric hydrocephalus, abnormalities in conventional DTI anisotropy measures have been associated with impaired neuromotor and cognitive development among PHH infants in other studies.^8, 9^ In a mixed cohort, Lean et al. demonstrated that IVH/PHH-related aberrant dMRI measures were associated with poorer motor, cognitive, and language scores at age 2 years compared to VPT infants. PHH infants specifically had white matter disruptions in the CST which related to poorer motor outcomes at age 2 years.^8^ These relationships likely result from disturbances in the complex sequence of events that underlie early white matter development due to IVH/PHH, engendering local and widespread effects through direct cellular mechanisms and disruption of subsequent cerebral development^34^. This sequence of events begins in the fetal VZ and SVZ zones, where neural stem cells (NSC) differentiate into oligodendrocytes, astrocytes, neurons, and mature ependymal cells^35^. In humans, these processes of neurogliogenesis continue into the newborn period, and thus are at risk precisely when IVH and PHH most commonly occur^36^. In the setting of IVH and PHH, differentiating NSCs are disrupted and often cleaved and released into CSF. Elevated levels of neurodevelopment-related proteins are differentially elevated in IVH/PHH compared with healthy infants^37^. Evidence for widespread VZ/SVZ disruption has also been reported in human postmortem tissues of preterm infants affected by IVH/PHH^38^. Collectively, the loss of progenitor cells and impaired neuroblast migration^39^, further impacted by pressure-related structural deformation, ischemia, inflammation, and other phenomena^40^, have brain-wide effects, affecting both cortical and subcortical regions and the development of white matter tracts^35^.

Studies of congenital hydrocephalus provide further evidence that hydrocephalus (independent of IVH) is associated motor and cognitive impairments that relate to dMRI measures. For example, Mangano et al. showed that infants with congenital (i.e., non-hemorrhagic) hydrocephalus who had aberrant fractional anisotropy and axial and radial diffusivity measures in the CC and CST scored poorly on ABAS-II and Bayley’s-III motor and cognitive scores compared with controls. While some of these effects persisted even up to one year following hydrocephalus surgery, marginal neurodevelopmental outcome improvements were observed.^9^

### Variability of white matter structure and development and their effects on dMRI findings

The variability in typical tract-specific patterns of white matter development must be taken into consideration when interpreting dMRI data. The data in this study were acquired at term equivalent postmenstrual age when some but not all white matter tracts are expected to have myelinated. Myelination begins and ends at variable ages among different white matter bundles. In general, myelination in the CST and OPRA start at 36- and 40-weeks’ gestation, respectively, whereas myelination in the CC does not begin until after two months postnatally^41^. Therefore, our findings are interpreted in the context of whether the assessed white matter is largely myelinated (CST and OPRA) or unmyelinated (CC). For myelinated tracts, FRD is largely taken to reflect myelin health. In unmyelinated tracts, it more likely reflects close fiber packing. In our study, FRD of the CC was unchanged in all three groups. In contrast, the myelinated CST demonstrated higher FRD in the PHH than IVH and VPT groups, suggesting myelin disruption/injury. For the myelinated OPRA, FRD was not different between groups suggesting that myelin was preserved. However, note also that FF was lower and FAD higher in the PHH group. These values are consistent with findings of transependymal flow injury reported in PHH infants^32, 42^ that may be associated with the tissue edema, hypercellularity, and inflammation.

### Ventricular size-dMRI correlation

Although ventricular size appears to be a risk factor for neurodevelopmental impairment in PHH, little is known regarding the link between ventricular size and white matter integrity. This underscores the importance of utilizing DBSI to evaluate the effects of ventricular size on white matter injury, providing a potential noninvasive biomarker for therapeutic responsiveness following surgical treatment of hydrocephalus. Larger ventricles were associated with axonal injury across all tracts, correlated negatively with fiber density in the CC and OPRA, and were associated with myelin injury in the CST, reflecting the literature suggesting a positive correlation between larger ventricular size and greater disruptions in white matter^32, 43^. Leveraging the unique results provided using DBSI, increases in ventricular size correlated with increased hypercellularity in the CC and edema in the CST and OPRA which may have been secondary to reactive cellular infiltration and inflammation, respectively, due to the detrimental effects of the expanding ventricles on white matter.

It has been shown that in PHH, the mechanical pressure from the large ventricles distorts PVWM fiber organization, decreases cerebral perfusion, and initiates an inflammatory cascade, all of which culminates in injury and axonal loss^40, 44^. In this cohort, the larger the ventricular size, the worse the DBSI metrics of axonal loss, edema, and hypercellularity, which all relate to neuroinflammation. These findings have critical clinical implications. Several studies have shown that progressive ventricular distension in PHH is associated with higher rates of neurological disability^9, 33^. Further, infants who undergo early surgical decompression based on increases in ventricular measurement have been shown to have better cognitive and neuromotor outcomes when compared to those who are treated relatively later based on clinical signs of increased ICP^45^. Thus, the findings of this study and the existing literature underscore the value of utilizing DBSI to evaluate the role of PHH and ventricular size – a readily modifiable factor – on white matter integrity, brain development, and neurodevelopmental disability associated with PHH.

### Histological correlates with DBSI

In this study, the DBSI finding of increased RF, representative of cellular infiltration, in the white matter of PHH infants was validated on H&E staining. The increased number of cells in white matter could be attributed, at least in part, to inflammatory processes such as gliosis and astrocytic proliferation, which have been shown to occur in the white matter of hydrocephalic human and animal tissues^38, 46^. Edema typically occurs in white matter in the setting of inflammation, which has been associated with the transependymal CSF flow observed in PHH^32, 42^. While variable amounts of edema were seen in the CC of the PHH infants as vacuolations on H&E staining, the DBSI correlate, NRF, did not differ between groups; this is presumably because NRF reflects interstitial or extracellular edema. Fiber loss, expressed as reduced FF, was demonstrated on immunohistochemistry as decreased synaptophysin, which suggested decreased neuropil and synaptic processes. White matter neuropil damage has been reported in both humans and experimental models of hydrocephalus and has been attributed to factors such as inflammation and fiber loss from mechanical compression of PVWM tracts by the expanding ventricles^47, 48^. It has been suggested that PVWM injury is preceded by or occurs in relation to damage of the lateral ventricular perimeter, which encompasses the VZ, SVZ, and immediately adjoining thin layer of white matter^32^. Similarly, in this study, our human PHH postmortem tissue analyses showed that the VZ/SVZ layer medial to the white matter injury were disrupted, demonstrating increased markers of inflammation including reactive astrocytosis and microglia/macrophage proliferation. These findings suggest that DBSI can provide a non-invasive neuroimaging modality for examining white matter cytoarchitecture at a granular level in PHH that directly relates to histology. However, further studies are required to examine its clinical utility.

### Limitations

A notable limitation of this study is the relatively small sample size of the PHH and IVH groups, which may have limited our ability to observe subtle DBSI differences between groups. There is also a potential for sex bias, as all 12 PHH patients were male. This reflects the fact that IVH tends to affect more males than females,^49^ and covarying for sex in our analyses did not affect the results. To minimize the effect of age on brain development, all scans were acquired cross-sectionally at term-equivalent postmenstrual age, which precludes assessment of the relationships between dMRI parameters and brain development over time.

## Conclusions

DBSI analyses of dMRI data collected at term equivalent age in very preterm infants demonstrated that PHH, and to a lesser magnitude IVH, were associated with PVWM disruption. While axonal fiber loss and hypercellularity were major underlying pathologies evident in all fiber tracts assessed, the OPRA and CST demonstrated evidence of edema, likely related to transependymal CSF migration. DBSI was able to uniquely distinguish direct effects on white matter from changes in the extracellular milieu. Representative histological analysis of postmortem tissue was consistent with the DBSI findings in the CC. Ventricular size correlated with DBSI measures in that larger ventricle size was associated with greater disruptions in the PVWM. These findings suggest that DBSI may provide a biomarker for clinical management and comparative effectiveness research in PHH to facilitate diagnosis and monitoring of therapeutic efficacy. In addition, novel insights into the pathophysiology of PHH gained through DBSI may enable investigation of new treatment strategies to minimize the developmental disability of this devastating disease. Future studies remain necessary to link these DBSI results to assessments of long-term developmental outcomes in an effort to elucidate the cellular mechanisms underlying impairment in high-risk VPT infants.

## Data Availability

All data supporting the results of this study are included in the manuscript

## ACKNOWLEDGEMENTS

We thank the IDDRC at Washington University for assistance with data collection; the children and their families for participating in the study, and we appreciate the support of our funding agencies. This work was supported by the Vanier Canada Graduate Scholarship [grant number 396212]; National Institutes of Health [grant numbers K02 NS089852, K23 NS075151, K23 MH105179, TL1 TR002344, P30 NS098577, R01 MH113570, R01 HD061619, R01 HD057098, R01 NS047592 and U01 EY025500]; Child Neurology Foundation; Cerebral Palsy International Research Foundation; The Dana Foundation; March of Dimes Prematurity Research Center at Washington University; The Doris Duke Charitable Foundation; Orion Pharma Research Foundation; and the Eunice Kennedy Shriver National Institute of Child Health & Human Development of the National Institutes of Health [grant number U54 HD087011]. None of the organizations listed had any role in the study design, data collection, data analysis, data interpretation, writing or decision to submit the report for publication. Our funding sources had no involvement in study design, data analysis, writing the report, or the decision to submit the article for publication.

## AUTHOR CONTRIBUTIONS

AMI, JJN, JPM, SD, LCR, HM, SP, SKS, DDL and CDS contributed to the conception and design of the study. AMI, JJN, JPM, SD, LCR, HM, HEB, DA, AG, SP, DM, YY and SKS were involved in data acquisition and analysis. AMI, JJN, JPM, SD, LCR, HM, HE, DA, SKS, DDL, DM, YY and CDS drafted significant portions of the manuscript and/or figures. All authors reviewed, edited and approved of the final draft.

